# The effect of CYP3A4 genetic polymorphisms on the pharmacokinetics of calcineurin inhibitors in renal transplant recipients: a systematic review and meta-analysis protocol

**DOI:** 10.1101/2021.03.13.21253531

**Authors:** Saeedeh Salehi, Fateme Abedini, Abbas Shahi, Shima Afzali, Simin Dashti-Khavidaki, Ali Akbar Amirzargar

**Affiliations:** Department of immunology, School of Medicine, Tehran University of Medical Sciences, Tehran, Iran; Student’s Scientific Research Center, Tehran University of Medical Sciences, Tehran, Iran; Department of immunology, School of Medicine, Iran University of Medical Sciences, Tehran, Iran; Nephrology Research Center, Tehran University of Medical Sciences, Tehran, Iran; Department of Clinical Pharmacy, School of Pharmacy, Tehran University of Medical Sciences, Tehran, Iran

## Abstract

**Background:** Calcineurin inhibitors (CNIs) are metabolized by CYP3A4. Polymorphisms in the CYP3A4 gene alter the activity of CYP3A4 protein and therefore affect the CNIs concentrations. Results of studies that investigated the association between CYP3A4 polymorphisms and CNIs pharmacokinetics are controversial. Therefore, this systematic review and meta-analysis will evaluate the effect of CYP3A4 polymorphisms on the pharmacokinetics of CNIs in renal transplant recipients.

**Method:** This protocol is developed according to the PRISMA-P guideline and registered in PROSPERO (CRD42020145219). The MeSH/Emtree terms of CYP3A4 polymorphisms and CNIs pharmacokinetics in PECO-based question will be obtained from the comprehensive literature search on PubMed/MEDLINE, Scopus, Web of Science, Embase, CENTRAL, and ProQuest without any language limitation from 1 January 1998 to 31 March 2021. Google Scholar search engine, registries, conference papers, and key journals will also be searched. Screening, selection, quality assessment, and data extraction will be performed by two independent reviewers. Statistical heterogeneity will be calculated by the Q Cochrane test and I^2^ statistic. Publication bias and sensitivity analysis will be evaluated by appropriate tests.

**Results and conclusion:** According to the meta-analysis of the aggregated data from the relevant primary studies, the association between CYP3A4 polymorphisms and CNIs pharmacokinetics will be reported, which possibly help the pharmacogenetic-guided dosing approach.

## 1. Introduction

The global burden of disease studies have reported that global deaths due to kidney failure were 1.2 million people in 2015, and the total number of deaths from this event increased by 32% from 2005 to 2015 [1]. Moreover, chronic kidney disease (CKD) has been introduced as the 13th and 17th leading cause of death in females and males, respectively [2]. The final stage of CKD is called end-stage renal disease (ESRD), and renal transplantation is the best treatment option for these patients. Based on the global observatory on donation and transplantation (GODT) report, 135860 solid organ transplantations were performed worldwide in 2016, which shows a 7.25% increase in comparison to 2015; among these, 89823 were renal transplants [3]. Graft survival greatly improved by using new surgical techniques and suitable immunosuppressive regimens. Current immunosuppressive protocols usually comprise a triple combination of immunosuppressive drugs such as calcineurin inhibitors (tacrolimus or cyclosporine A), anti-proliferative agents, and glucocorticoids with or without induction therapy [4, 5]. Tacrolimus and cyclosporine were firstly approved for liver and kidney transplantation, respectively. However, because of their acceptable efficacy, they expanded as the first-line component of the immunosuppressive regimen for the other solid organ transplantations [4]. Unfortunately, the pharmacokinetics of CNIs considerably differ between individuals [6-9]. Multiple physiological, environmental, and genetic factors are identified which influence the pharmacokinetic characteristics of CNIs, including the recipient’s age, sex, body weight, ethnicity, concurrent medications (azole antifungal drugs, macrolide antibiotics, non-dihydropyridine calcium channel blockers, rifampin, carbamazepine, phenytoin, dietary supplements, etc.), pharmaceutical formulation (brand or generic), post-transplantation period, food ingredients, gastrointestinal complications (constipation or diarrhea), ascites, hepatic function and some recipient’s genetic factors [10-14]. Genetic makeup can cause a 20-95% variation in drug metabolism [15]; therefore, recent pharmacogenetic studies focused on genetic variations such as single nucleotide polymorphism (SNP), which may affect response to drugs [15]. One of the highly polymorphic genes which influences drug metabolism is cytochrome P450 (CYP). In the human genome, CYP has approximately 57 active genes that encoded functional enzymes and classified into 18 families and 44 subfamilies. Members of CYP1-3 families, especially CYP3A4 and CYP3A5, are involved in drug metabolism. CYP3A4 is estimated to account for about 50% of all CYP-mediated drug metabolism [6, 16]. However, variations of CYP3A4 expression levels between different individuals cause different metabolizing activities of this enzyme. This difference is at least partly due to polymorphism in the enzyme-encoding genes that can lead to decreased (CYP3A4*22 and CYP3A*26 SNPs) or increased (CYP3A4*1B, CYP3A4* 1G, and CYP3A4*18 SNPs) enzyme activity [6]. Therefore, this factor can be regarded as a drug dosing determinant.

Several primary studies have investigated the relation of CYP3A SNPs and pharmacokinetics of CNIs in solid organ transplant recipients, but the results are controversial [17-24]. Furthermore, few meta-analysis studies have appraised the effects of CYP3A4 SNPs on the pharmacokinetics of tacrolimus [25] and cyclosporine A [26, 27], which have not yielded conclusive results in renal transplant recipients. On the other hand, recently, a pharmacogenetic-guided CNIs dosing approach has been considered. This strategy facilitates personalized medicine, in which genetic makeups of patients determine optimum doses of immunosuppressive drugs and proper drug combinations. To the best of our knowledge, no systematic review and meta-analysis study has investigated the influence of all effective CYP3A4 SNPs on pharmacokinetic characteristics of both CNIs (tacrolimus and cyclosporine A) in kidney transplant recipients. Consequently, we decided to perform this systematic review and meta-analysis on all relevant primary studies to explore the impact of CYP3A4 SNPs on the pharmacokinetics characteristics of both available CNIs.

## 2. Method and analysis

### 2.1. Study design

This protocol is developed according to the Preferred Reporting Items for Systematic reviews and Meta-Analyses guidelines for protocols (PRISMA-P) [28] (<u>Appendix 1</u>). A Flow diagram will be drawn to display the details of excluded studies and their reasons (figure 1). This research project was prospectively registered in the International Prospective Register of Systematic Reviews (PROSPERO) with the CRD42020145219 registration number.

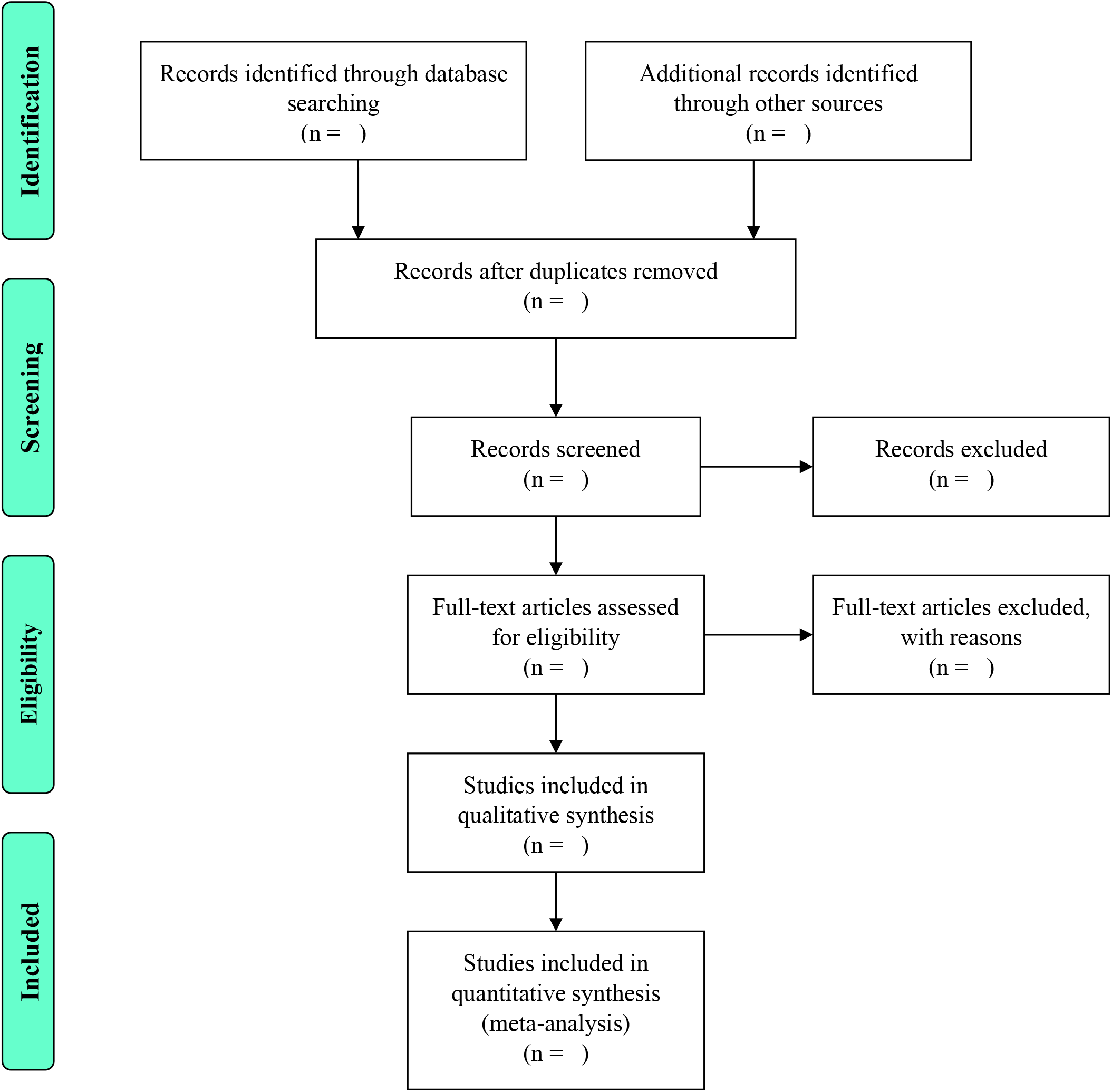
PRISMA flow diagram.

### 2.2. Eligibility criteria

#### 2.2.1. Types of studies

a. All types of observational studies, including case-control, cohort (retrospective or prospective), cross-sectional studies that are designed to examine the impact of at least one of the CYP3A4 gene polymorphisms on at least one type of CNIs pharmacokinetics in renal transplant recipients, will be included.
b. Relevant baseline data of clinical trials that are registered in registry systems such as ClinicalTrials.gov will be included.
c. Case reports, case series, letters, review articles, and animal studies will be excluded.

#### 2.2.2. Types of participants

a. In this study, all age groups (pediatric, adult, and elderly) and both sex groups who have received renal transplantation will be included. If a primary study is done on at least one age or sex group, it will be included, too.
b. There is no limitation regarding the recipient’s ethnicity for inclusion in this review.
c. Renal transplantations can be received from the living or deceased donor. Kidney transplantation can be done de novo (before dialysis treatment) or secondary (after treating by dialysis for a while).
d. Patients have not taken medications interacting with CNIs.
e. Participants have not the underlying disease which affects CNIs pharmacokinetics.

#### 2.2.3 Exposure

a. In the immunosuppressive regimen of the patients, one type of brand or generic formulations of tacrolimus or cyclosporine A has been used.
b. This study does not have any limitation for a method of measuring the concentration of CNIs, i.e., primary studies with any measurement method such as CMIA, MEIA, ACMIA, CLIA, FPIA, LC-MS/MS, EMIT, RIA, CEDIA, HPLC, mass spectrometry, etc. and these measurements can be done on any biologic specimen (blood, plasma, serum).

##### 2.2.4 Outcome

a. For tacrolimus: Dose-adjusted trough concentration (C_0_/D) should have been reported or could be estimated by the corresponding 24-hour dose.
b. For cyclosporine A: Dose-adjusted trough concentration (C0/D) or dose-corrected peak concentration (C_2_/D) should have been reported or could be estimated by the corresponding 24-hour dose.

### 2.3. Search strategy

#### 2.3.1. Official databases

The comprehensive literature search will be conducted on electronic databases such as PubMed/MEDLINE, Scopus, Web of Science, Embase, and CENTRAL without any language limitation from 1 January 1998 (Because the first peer-reviewed CYP3A4 SNP discovered in this year) to 31 March 2021 (the search time interval was extended). Non-English papers will be translated by free online translators. Search syntax will be developed in PubMed, and after achieving sufficient search syntax, it will be modified as required for other databases. Related terms with E (CYP3A4 SNPs) and O (CNIs pharmacokinetics) in PECO-based question will be obtained from the three-phase method, as well as MeSH/Emtree terms and free text method.

#### 2.3.2. Other sources

According to the recommendation of the “Cochrane Handbook for Systematic Reviews of Interventions” [29] and “Institute of medicine standards for the systematic review of comparative effectiveness research “ [30], we will include gray literature in this research study. An electronic search of Google Scholar search engine and registry systems and conference papers (indexed in ProQuest, Scopus, Web of Science, and Embase) will also be performed. Two key journals based on the Scopus report, and as well as reference lists of included primary studies, will be searched manually for additional studies.

### 2.4. Data management in the specific condition

In the case of unavailable necessary data in included primary studies, we will contact the corresponding author in the intervals of two weeks for three times. If we do not catch any response, we will exclude that paper. If one study has multiple publications, we will use its valid and complete version. For graphs without any data in the text, we will utilize the WebPlotDigitizer-4.2 to extract the required data [31].

### 2.5. Screening and selection

Related papers from any databases will be transferred to the EndNote software version X 8 and duplicated papers will be removed from the library. Two independent reviewers will be screened the title and abstract of retrieved search results and then evaluate full texts of the remaining papers for identifying papers regarding the eligibility criteria mentioned above by the same two reviewers. Disagreements will be resolved through consensus between the authors, and when consensus is not achieved, a third reviewer will act as a referee, and if it’s not possible, a third assessor will act as a referee.

### 2.6. Data extraction

Collection and data analysis for this study will be performed according to the Preferred Reporting Items for Systematic Reviews and Meta-Analyses (PRISMA) checklist and statement [32].

Data extraction will be done by two reviewers. Disagreements will be resolved by consensus, and when consensus is not achieved, a third reviewer will act as an arbitrator. We will design the data abstraction form for the reviewers. Extracted data from included primary studies will be:

#### 2.6.1. Study characteristics

The first author, year of publication, study design, follow-up duration (average time after transplantation), sample size, and location.

#### 2.6.2. Participants characteristics

Recipient’s age, sex, weight, body mass index (BMI), ethnicity, functional parameters such as estimated glomerular filtration rate (eGFR), hematocrit (pre-transplantation, post-transplantation), donor type (deceased, living), immunosuppressive regimen, and induction therapy.

2.6.3. Exposure characteristics

Type of CNI (tacrolimus or cyclosporine A), type of formulation (brand or generic), the measurement method of CNI concentration (CMIA, MEIA, ACMIA, CLIA, FPIA, LC-MS/MS, EMIT, RIA, CEDIA, HPLC, mass spectrometry), biological specimen (blood, serum, plasma), CNI trough concentration, CNI dose-adjusted trough concentration (C_0_/D), and cyclosporine A dose-adjusted peak concentration (C_2_/D).

#### 2.6.4. Outcome

DNA extraction method, SNP types (CYP3A4*1B, CYP3A4*1G, CYP3A4*18, CYP3A4*22), and allele/genotype frequency.

### 2.7. Risk of bias assessment

Quality assessment of the selected primary studies will be accomplished by two authors independently according to the New-castle Ottawa scale (NOS), which has three options: selection (4 items), comparability (1 item), and outcome (3 items) [33]. The question which is received the star (“a,” and in some questions “a” and “b”) will be calculated in the final score. Subsequently, included studies will be classified into three levels: good, fair, and poor quality. Discrepancies will be resolved by consensus.

#### 2.8. Assessment of statistical heterogeneity

Statistical heterogeneity will be assessed by the Q Cochrane test (*χ2*) and severity of heterogeneity evaluated by I-squared (I^2^) [34]. I^2^ value will be defined as the following guide [35]:

1. 0-40%: mild heterogeneity
2. 30-60%: moderate heterogeneity
3. 50-90%: severe heterogeneity
4. 75-100%: highly severe heterogeneity.

When there is a substantial heterogeneity (i.e., I^2^>50%), either subgroup analysis or meta-regression will be conducted to determine the possible source of heterogeneity based on these characteristics: Age group, ethnicity, type of CNI (tacrolimus or cyclosporine A, brand or generic), measurement method of CNI concentration, post-transplantation time, DNA extraction method, SNP types (CYP3A4*1B, CYP3A4*1G, CYP3A4*18, CYP3A4*22), genotyping detection method, C_0_/D, and C_2_/D.

### 2.9. Assessment of publication bias

The publication bias will be evaluated by the “Funnel plot” [36], “Begg’s test”, “Egger’s test and plot” [37], and “trim and fill method” [38, 39].

### 2.10. Assessment of sensitivity analysis

We will perform a sensitivity analysis by the one-out remove (leave-one-out) method to assess the validity of the pooled estimates. In summary, by excluding one primary study and repeating the analysis, its influence on the overall estimate will be conducted.

### 2.11. Statistical analysis (meta-analysis or meta-synthesis)

Data analyzing will be performed by Stata software version 14 (StataCorp. 2015. Stata Statistical Software: Release 14. College Station, TX: StataCorp LP). The meta-analysis will be done to evaluate the effect of CYP3A4 SNPs on the pharmacokinetics of CNIs in renal transplant recipients. If the meta-analysis is not possible, Meta-synthesis may be applied. Key measures of meta-analysis, including sample size, mean difference, and the standardized mean difference, will be combined by a fixed or random-effect model (FEM, REM). The graphical presentation of data will be conducted by the forest plot. A *P*< 0.05 will be defined as statistically significant.

#### 2.12. Ethics and dissemination

As this study is the analysis of primary researches, no ethics approval will be required. We will publish the final results of this systematic review and meta-analysis in a peer-review journal. Also, it may present in the international or national congresses.

## 3. Results

This systematic review and meta-analysis is ongoing, and the process is continuing. According to the meta-analysis of the aggregated data from the relevant primary studies, the association between CYP3A4 polymorphisms and CNIs pharmacokinetics will be reported, which possibly help the pharmacogenetic-guided dosing approach.

## 4. Discussion

CYP3A4 is a member of the cytochrome P450 monooxygenase enzymes family prominently expressed in the small intestine, liver, and rarely kidney and lung of all individuals and efficiently contributes to metabolizing of cyclosporine A and tacrolimus via oxidation-reduction reactions. Genetic polymorphisms in the CYP3A4 gene can decrease or increase the CNI-metabolizing activity of this enzyme. Therefore, the determination of these polymorphisms in a transplanted patient has the potential to identify patients who are at graft rejection risk due to inappropriate immunosuppressive levels or those who may be more prone to side effects. Ultimately, genotyping may lead to further personalization of immunosuppressive therapy for transplant recipients. The reviewing of results will inform clinicians and guideline developers about the best available evidence. Also, these results may show which CNIs, cyclosporine A or tacrolimus, are more effective for renal transplant recipients with a specific CYP3A4 polymorphism. This protocol defines our objectives and method for the meta-analysis of the primary observational studies. We will search all bibliographic databases and potential gray literature sources with no language restriction to perform a better systematic review and meta-analysis. Our study has several strengths such as, adding two more important database (Scopus, ProQuest), conference papers, and thesis in comparison to other similar studies; also primary studies that were published between October 2017 (search termination time of last relevant meta-analysis study) and 31 March 2021 will be added to this study. Moreover, non-English papers will be added to this study, and publication bias of primary studies will be evaluated at least by two methods. We will assess the effect of all functional polymorphisms of CYP3A4 on the pharmacokinetics of both types of calcineurin inhibitors. Eventually, we expect that our study significantly advances the knowledge about the effect of specific CYP3A4 polymorphisms on CNIs dose and concentration.

## Data Availability

This study is ongoing, and its findings will be available from the corresponding author upon request.

## Author contribution

SS, AS, and FA prepared the primary draft of the protocol under the guidance of SDK. SS and SA will develop sufficient search syntax and modify it as required for other databases. Screening and selection will be done by SS and FA under the supervision of AAA. Quality assessment of the included studies will be done by SS and FA under the guidance of AAA. Data extraction will be performed by AS and SA under the superintendence of SDK. All listed authors will critically revise the final version of the manuscript before the publication.

## Acknowledgment

The authors acknowledge the Department of Immunology, School of Medicine, Tehran University of Medical Science, Tehran, Iran, for their assistance.

## Conflicts of interest

None.

## Notes

### Competing Interest Statement

The authors have declared no competing interest.

### Clinical Protocols

https://www.crd.york.ac.uk/prospero/display_record.php?RecordID=145219

### Author Declarations

This research is a secondary study, and ethics committee approval is not necessary.

